# Diabetes Polygenic Scores Predict Glycemic Indices and Insulin Use in Individuals with Atypical Diabetes

**DOI:** 10.64898/2026.10.01.26364299

**Authors:** Steven D. Gage, Raymond J. Kreienkamp, Aaron J. Deutsch, Eric J. Richards, Liana K. Billings, Sara J. Cromer, Siri Atma W. Greeley, Irl B. Hirsch, Alicia Huerta-Chagoya, Steven E. Kahn, Klara R. Klein, Josep M. Mercader, Neda Rasouli, Marjan Rezaei, Maria J. Redondo, Ashok Balasubramanyam, Jose C. Florez, Louis H. Philipson, Jason Flannick, Miriam S. Udler, RADIANT Study Group

## Abstract

**Aims/Hypothesis:** Atypical forms of diabetes often provide diagnostic challenges and are suboptimally treated. We sought to determine whether type 1 and 2 diabetes polygenic scores (PS) would help improve classification of islet autoantibody-negative (IAb-) atypical diabetes cases.

**Methods:** We implemented type 1 diabetes PS (T1D PS) and type 2 diabetes PS (T2D PS) in 309 IAb-individuals’ genome sequencing from the Rare and Atypical Diabetes Network (RADIANT), the largest and most comprehensively phenotyped collection of unsolved and atypical diabetes cases. We performed regression analyses to assess associations between PS, clinical characteristics, and glycemic measures.

**Results:** Both T1D and T2D PS were significantly higher in RADIANT than in ancestry-matched controls from UK Biobank. Both PS were associated with C-peptide measures throughout participant oral glucose tolerance tests (OGTT), although in opposite directions; T1D PS was associated with lower C-peptide measures, while T2D PS was associated with higher measures. Participants with T1D PS above a previously established optimal PS value for differentiating cases and controls (T1D PS GRS2>12.88) had 13-fold higher odds of both insulin deficiency on OGTT and use of both basal and bolus insulin therapy.

**Conclusions/Interpretation:** Individuals in RADIANT with ambiguous diabetes subtypes displayed genetic enrichment for both type 1 and type 2 diabetes, despite ascertainment to remove standard type 1 and type 2 diabetes. T1D PS identified individuals with an unrecognized “type 1 diabetes-<u>like</u>” phenotype with insulin deficiency and need for intensive insulin in the absence of islet autoantibodies. PS may help identify individuals with atypical diabetes who will ultimately benefit from insulin therapy.

## Introduction

Diabetes is classified into subtypes, including type 1 diabetes and type 2 diabetes, to capture more precisely the specific diabetes phenotype and etiology [1]. These classifications simplify the disease to a commonly accepted treatment paradigm and anticipated clinical trajectory. Yet, some individuals do not clearly match the classic diabetes subtypes and have “atypical” diabetes [2]. These individuals can be clinically challenging, as the optimal treatment may not be readily evident. Therefore, new tools are needed to aid clinicians in these scenarios.

Beyond molecular genetic testing for monogenic diabetes, genetics has not been widely utilized in diabetes clinical care. However, as diabetes has a strong genetic basis [3, 4], genetic information might aid characterization of these atypical cases. Polygenic scores (PS), which sum the risk alleles associated with a disease across the genome, estimate an aggregate risk along a continuous scale and can identify individuals with high genetic risk for type 1 or type 2 diabetes. Type 1 diabetes PS (T1D PS) have high discriminatory capacity, with an area under the receiver operating characteristic curve (AUC) >0.9 for separating type 1 diabetes from all other individuals [5]. In fact, T1D PS can distinguish type 1 diabetes from type 2 diabetes [6], monogenic diabetes [7], and monogenic autoimmunity [8]. Applications for T1D PS in clinical practice are increasingly being recognized [9]. Type 2 diabetes PS (T2D PS) have lower differentiating capacity, with AUC ∼0.7, although they may still provide some clinical benefit [10–12].

The goal of the Rare and Atypical Diabetes Network (RADIANT) is to characterize diabetes cases that do not clearly match classic diabetes subtypes [13]. It is the largest study dedicated to the collection of atypical diabetes cases to date. An adjudication committee comprised of experienced diabetologists and geneticists reviews all clinical data and laboratory measures, including islet autoantibody testing, to rigorously exclude classic type 1, type 2, or monogenic diabetes. RADIANT has performed genome sequencing and clinical phenotyping in >300 islet autoantibody-negative (IAb-) participants.

The extent to which T1D PS and T2D PS might provide phenotypic clarity in a cohort of real-world atypical diabetes cases is currently unclear. Given the need for additional clinical tools in atypical diabetes, we implemented diabetes PS in individuals with atypical diabetes in RADIANT and assessed whether PS provide insights into the genetic architecture of an atypical diabetes population.

## Methods

### Study Population

The Rare and Atypical DIAbetes NeTwork (RADIANT) seeks to identify and characterize patients with atypical diabetes and define novel subtypes of atypical diabetes [13]. It is the largest and most comprehensively phenotyped collection of atypical diabetes cases to date. Details of the RADIANT study protocol can be found elsewhere [13]. Briefly, individuals with diabetes but phenotypes inconsistent with classic type 1 or type 2 diabetes were referred by their clinical provider or self-enrolled. Individuals completed a comprehensive questionnaire detailing their medical history (stage 1). Participants whose phenotype was inconsistent with typical diabetes were selected by an adjudication committee comprised of expert diabetologists and geneticists. Subsequently, individuals underwent genome sequencing (GS, stage 2) and comprehensive phenotyping (stage 3), including laboratory studies and an oral glucose tolerance test (OGTT). The current study includes RADIANT participants with GS and comprehensive phenotyping completed by June 30, 2025, with negative islet autoantibodies and complete medication information (n=309). The RADIANT protocol was approved by the University of Utah Institutional Review Board.

For comparison, we analyzed array-based genetic data and medical record-based phenotype data for up to ∼487,000 participants from the UK Biobank (UKBB) [14]. Individuals in the UKBB were classified as having type 1 diabetes [5], type 2 diabetes [15], or no diabetes [15] using previously defined medical record-based algorithms. Analysis of the UK Biobank was conducted under application #42614. Data analysis for both studies was approved by the Mass General Brigham Institutional Review Board (Boston, MA).

### Polygenic Score Generation

RADIANT GS was conducted by the Laboratory for Molecular Medicine (LMM) at Mass General Brigham. GS participant data were aligned to GRCh38 (sequenced by LMM/Broad Institute of MIT and Harvard) and used to create joint variant calling format files with HaplotypeCaller 3.5 (Whole Genome Analysis Pipeline / WARP pipelines / Broad Data Sciences Platform) on the Terra.bio platform.

A T1D PS (T1D GRS2 [5]) and T2D PS (derived from Suzuki et al. [11]) were calculated from RADIANT GS data. As negative controls, an attention-deficit/hyperactivity disorder (ADHD) PS [16] and a height PS [17] were also calculated. Scoring alleles were counted and scored with PLINK2. The T1D PS was computed with 67 genome-wide significant variants, accounting for interactions between human leukocyte antigen (HLA) alleles [5]. One T1D PS scoring allele (rs3129197) was not called in RADIANT. A proxy allele (rs3129198) was identified using LDlink [18]; the alleles have a linkage disequilibrium R^2^ = 1.0 and D’ = 1.0 across all ancestries in the 1000 Genomes Project [19]. The T2D PS includes over 1,000,000 genetic variants (including those that do not reach genome-wide significance) from a recent multi-ancestry genome-wide association study of type 2 diabetes [11]. PS were also calculated in UKBB participants using array-based genotype data, imputed using TOPMED [20]. Population means between RADIANT and UKBB participants of European ancestry were compared directly using Welch’s t-test without adjustment.

### Genetically Inferred Ancestry

Principal component (PC) analysis was performed by the Pan-UK Biobank team to infer genetic ancestry in the UKBB [21]. Continental-level participant ancestry was inferred by projecting PCs onto a reference panel constructed from the Human Genome Diversity Project and the 1000 Genomes Project. Individuals were classified into one of five continental-level ancestry groups: Admixed American, African, East Asian, European, or South Asian. A similar method was used to infer genetic ancestry in RADIANT, after performing PC analysis using FlashPCA2 [22]. In RADIANT, due to low sample size in the East Asian and South Asian groups, these categories were combined into a single group (Asian) for subsequent generation of intra-ancestry PCs for statistical modeling.

### Statistical Analyses and Presentation

All statistical analyses were performed using R 4.3.3 [23] and specified packages, and all plots were generated using “ggplot2” 3.5.1 [24].

For the primary analyses, T1D PS and T2D PS were modeled as continuous variables using linear regression. As a sensitivity analysis, we compared linear and generalized additive models to assess whether allowing for nonlinearity improved the fit of the PS– C-peptide relationship. Model comparisons using Akaike information criterion and analysis of variance provided no evidence of improved fit with the more flexible model.

As a secondary analysis, T1D PS was modeled as a binary variable using logistic regression. For the binary variable, individuals were classified as having a “high” T1D PS if their T1D PS was greater than 12.88, the PS value previously established with maximum Youden’s index (YI_max_) to separate type 1 diabetes cases from controls in the UKBB [5].

All regression analyses used the same standard set of covariates (sex, age, duration, BMI, and the first 10 intra-ancestry PCs) unless otherwise explicitly stated. Regression models were performed across each of the four continental ancestry groups and meta-analyzed in a fixed-effects model. If ancestry-specific modeling could not be performed due to small sample size, then analyses were limited to European ancestry, as explicitly stated. Testing for linear trend (p-trend) was performed for diabetes duration categories with coeftest (“lmtest”, 0.9-40 [25]) using standard covariates.

For phenotype analyses, C-peptide levels were obtained from OGTT. The C-peptide index was calculated as follows [26]:

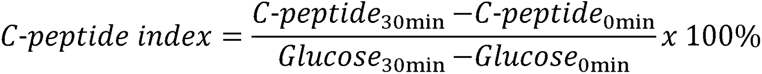

## Results

### Both T1D and T2D PS contribute to the genetic architecture of atypical diabetes

The RADIANT study includes an array of atypical diabetes presentations across the lifespan, with phenotypes deemed by expert clinicians to not match typical type 1 or type 2 diabetes (Table 1). To benchmark RADIANT to more typical diabetes phenotypes, we compared T2D PS and T1D PS in RADIANT to participants in the UKBB with type 1 or type 2 diabetes, as well as controls without diabetes. For this analysis we focused on participants of European genetic ancestry (207 in RADIANT and 285,728 in the UKBB) to avoid residual confounding from population stratification. We confirmed validity of comparing PS between these distinct datasets by calculating PS for two control phenotypes, ADHD and height, and demonstrating no significant differences between populations (Supplemental Fig. 1).

**Table 1.** RADIANT cohort characteristics. Clinical measures and prescription status were determined during Stage 3 site visit. Genetic ancestry was inferred by principal component (PC) analysis. ^†^ In order to maintain sufficient sample size, regression models using ancestry PC cluster East Asian and South Asian together.

| Clinical Measures | Median (1Q, 3Q) |
| --- | --- |
| Number of Participants | 309 |
| Age at Stage 3 visit (years) | 47 (35, 60) |
| Age at Diabetes Diagnosis (years) | 31.5 (20.8, 44.2) |
| Diabetes Duration (years) | 10 (4, 20) |
| BMI (kg/m<sup>2</sup>) | 25.6 (21.6, 27.3) |
| HbA1c (mmol/mol) | 52 mmol/mol (42, 58) |
| HbA1c (%) | 6.88% (6.0%, 7.45%) |
| Sex |  |
| *Female* | 167 (54.0%) |
| *Male* | 142 (46.0%) |
| Inferred Genetic Ancestry |  |
| *Admixed American* | 28 (9.1%) |
| *African* | 34 (11.0%) |
| *East Asian*<sup>†</sup> | 17 (5.5%) |
| *European* | 207 (67.0%) |
| *South Asian*<sup>†</sup> | 23 (7.4%) |
| Diabetes (DM) Medication Status |  |
| Any Diabetes Medication | 296 (95.8%) |
| Non-insulin Diabetes Medication | 199 (64.4%) |
| Insulin Prescription | 181 (58.6%) |
| Basal Insulin Only | 26 (8.4%) |
| Basal and Bolus Insulin | 138 (44.7%) |

We found that the mean T2D PS of RADIANT participants was significantly higher than that of controls or type 1 diabetes cases in the UKBB, and it was statistically indistinguishable from UKBB type 2 diabetes cases (Fig. 1). We next assessed the performance of the T1D PS. The mean T1D PS in RADIANT was significantly lower than the type 1 diabetes group in the UKBB, but significantly higher than the control and type 2 diabetes groups. These results indicated that despite efforts to exclude individuals with classic type 1 or type 2 diabetes from RADIANT, there was still evidence of increased polygenic risk for both type 1 and type 2 diabetes among RADIANT participants with IAb-atypical diabetes.

**Figure 1.**
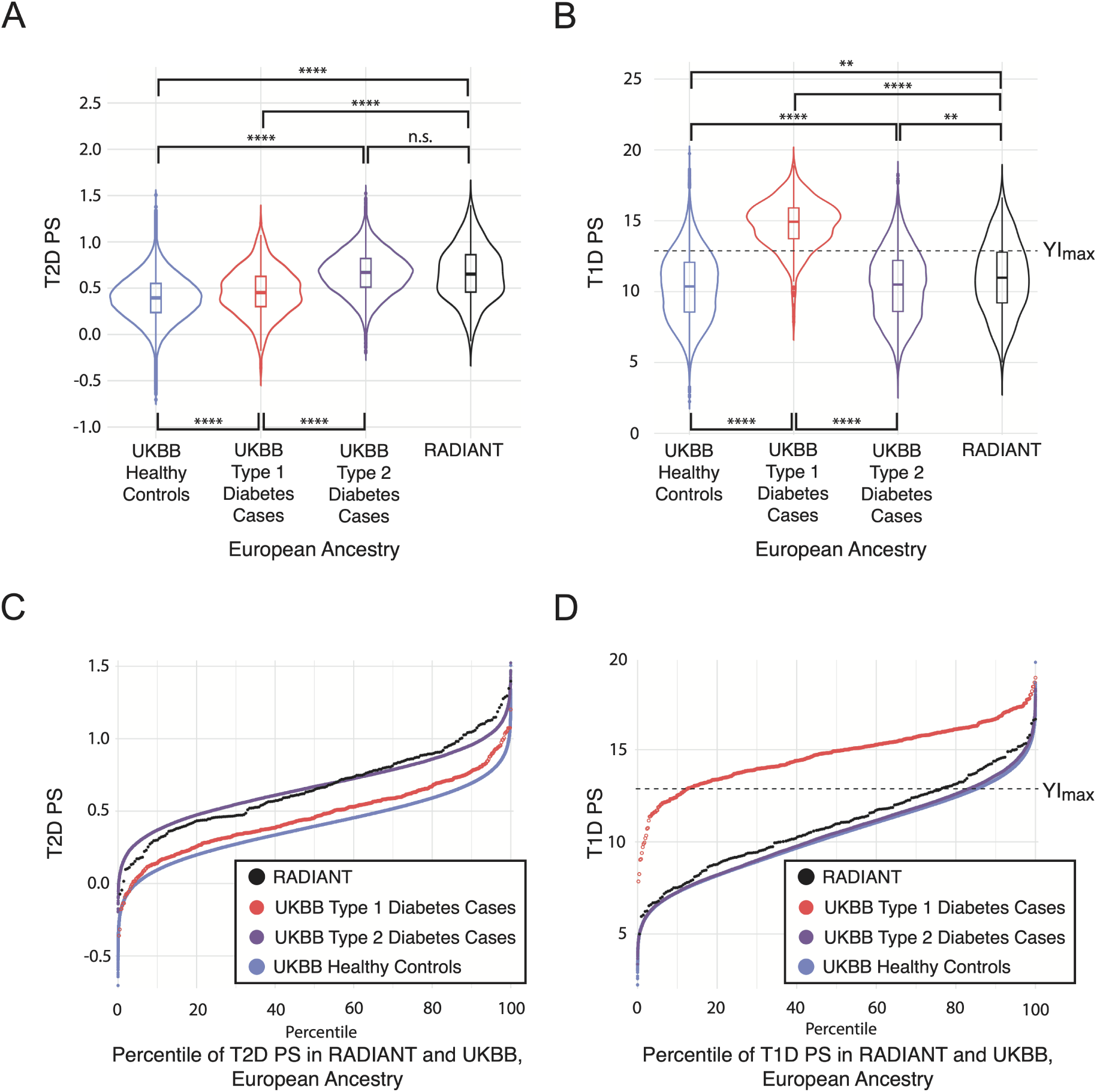
Distribution of T1D PS and T2D PS among the UK Biobank (UKBB) and RADIANT Participants of European Ancestry. **A & B** Violin plots of T2D PS (**A**) or T1D PS (**B**) distributions for UKBB control participants (blue), UKBB type 1 diabetes cases (red), UKBB type 2 diabetes cases (purple), and RADIANT participants (black), all of European ancestry. For T1D PS, the value with maximum Youden’s Index (YI_max_) is labeled with a dashed line, T1D PS=12.88. Significant differences in means were determined by Welch’s t-test; n.s., *p*>0.05; **, *p*<0.01; ****, *p*<0.0001. **<u>C & D</u>**, Individual participant T2D PS (**C**) or T1D PS (**D**) sorted by rank. Black circles, RADIANT participants; blue circles, UKBB control participants; red circles, UKBB type 1 diabetes cases; purple circles, type 2 diabetes cases, all of European ancestry.

### T1D PS predicts β-cell function in IAb-atypical diabetes

We assessed how diabetes PS predicted pancreatic β-cell function in RADIANT. In a meta-analysis across continental ancestry groups, increased T1D PS was associated with a lower C-peptide index and lower C-peptide secretion at all OGTT timepoints (Fig. 2); for instance, a one standard deviation (SD) increase in T1D PS was associated with a 0.16 nmol/l lower fasting C-peptide (*p*=1.0×10^-8^) and 0.49 nmol/l lower 120-min C-peptide (*p*=4.7×10^-7^). Conversely, a one-SD increase in T2D PS was associated with increased C-peptide secretion at all OGTT timepoints, including 0.12 nmol/l higher fasting C-peptide (*p*=4.3×10^-5^) and 0.37 nmol/l higher 120-min C-peptide (*p*=2.8×10^-4^). C-peptide index also significantly decreased per SD T1D PS (*p*=6.8×10^-4^), but not per SD T2D PS (*p*=0.14).

**Figure 2.**
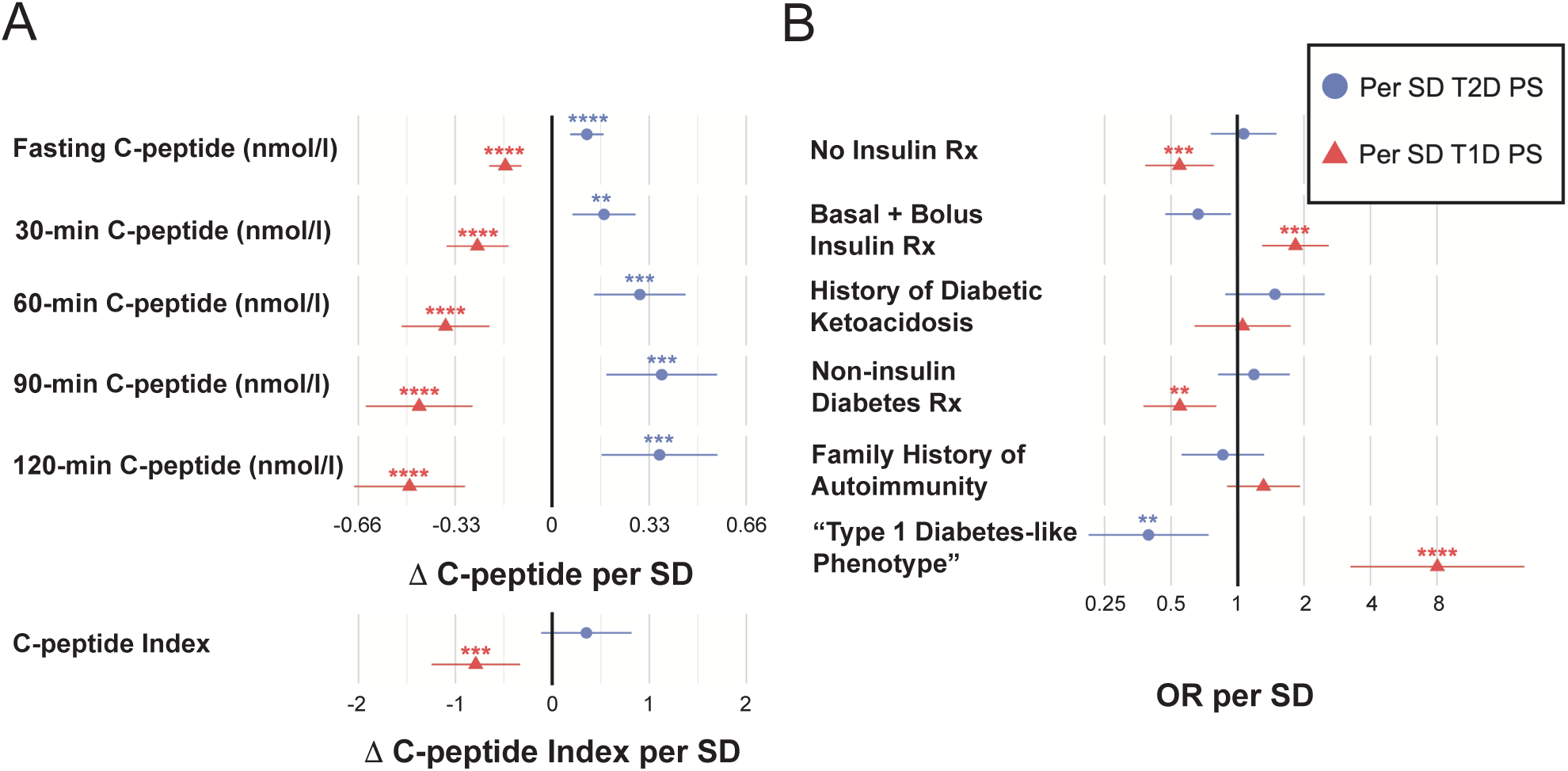
Relationship between PS, C-peptide levels, and clinical history. Association between C-peptide measurements from oral glucose tolerance tests (**A**) or participant history (**B**) with T1D PS (red triangles) or T2D PS (blue circles). Values were obtained from linear regression models (**A**) or logistic regression models (**B**), controlling for standard covariates and meta-analyzing across ancestries. SD, standard deviation; OR, odds ratio. Significance surpassing Bonferroni correction is labeled with asterisks; *p*<0.01, **; *p*<0.001, ***; *p*<0.0001, ****.

We then assessed T1D and T2D PS associations with phenotypic features of diabetes. In our covariate-adjusted, ancestry meta-analyzed logistic models, each one-SD increase in the T1D PS reduced the likelihood of “no insulin” use (OR=0.55, *p*=8.8×10^-4^) and “non-insulin diabetes medication” use (OR=0.55, *p*=1.9×10^-3^). We also assessed individuals we described as having a “type 1 diabetes-<u>like</u>” phenotype, who required both basal and bolus insulin, had a fasting C-peptide <0.20 nmol/l, and had a peak C-peptide <0.6 nmol/l [1]. An adjusted, ancestry-meta-analyzed one-SD increase in T1D PS was associated with increased odds for requiring both basal and bolus insulin, whether with a pump or injections (OR=1.83, *p*=6.5×10^-4^), and having a “type 1 diabetes-<u>like</u>” phenotype (OR=8.0, *p*=6.6×10^-6^), while a one-SD increase in T2D PS reduced the odds of requiring basal and bolus insulin (OR=0.66, *p*=1.8×10^-2^) and “type 1 diabetes-<u>like</u>” phenotype (OR=0.39, *p*=3.6×10^-3^). Neither PS was associated with history of diabetic ketoacidosis (DKA) nor family history of autoimmunity based on history available.

Because T1D PS and T2D PS were each associated with clinical phenotypes, we assessed whether a combined model that simultaneously accounted for T1D PS and T2D PS would yield better prediction of clinical variables. However, the adjusted combined model did not yield substantially better results compared to a model with T2D PS or T1D PS alone (Supplemental Fig. 2). We explored whether this was due to asymmetric polygenic contributions by determining the incremental value of each PS using nested models, comparing models with covariates alone, each PS individually, and both PS together. For all C-peptide measures, we observed a larger contribution from T1D PS than T2D PS (N-weighted marginal ΔR²: T1D PS 0.043–0.100 versus T2D PS 0.011–0.058). In the combined model, T1D PS had significant independent contribution to most clinical characteristics, unlike T2D PS.

### High T1D PS delineates a subgroup with reduced β-cell function

Next, we assessed whether the T1D PS could meaningfully stratify participants, focusing on T1D PS due to its reported strong differentiating capacity for distinguishing type 1 diabetes cases from all other individuals at a threshold PS value of 12.88 (Youden’s index YI_max_) [5]. We compared clinical and glycemic characteristics in individuals with T1D PS above this threshold (high T1D PS) to everyone else using a multi-ancestry adjusted model (Supplemental Table 1). Notably, participants with high T1D PS had markedly higher odds of requiring basal and bolus insulin (whether by pump or injections; OR=6.3, *p*=4.2×10^-5^) and their C-peptide levels during OGTT were also significantly lower across all time points (Supplemental Fig. 3).

We also considered whether the association with lower C-peptide levels in these individuals with high T1D PS might be modified by diabetes duration (Fig. 3, Supplemental Fig. 4), as C-peptide levels are known to wane over time [27]. After stratifying by windows of diabetes duration (≤2 years, >2 and ≤10 years, >10 years), mean C-peptide index values in the high T1D PS group were largely unchanged over duration periods, whereas the values significantly declined over time in participants without a high T1D PS (p-trend=6.5×10^-3^). Similarly, 120-min C-peptide values did not significantly change over time in the high T1D PS group. In the non-elevated T1D PS group, the downward trend in mean 120-min C-peptide values across duration periods did not reach statistical significance, although values were overall elevated compared to the high T1D PS group, with a significant difference detectable in intermediate duration period (*p*=4.1×10^-5^).

**Figure 3.**
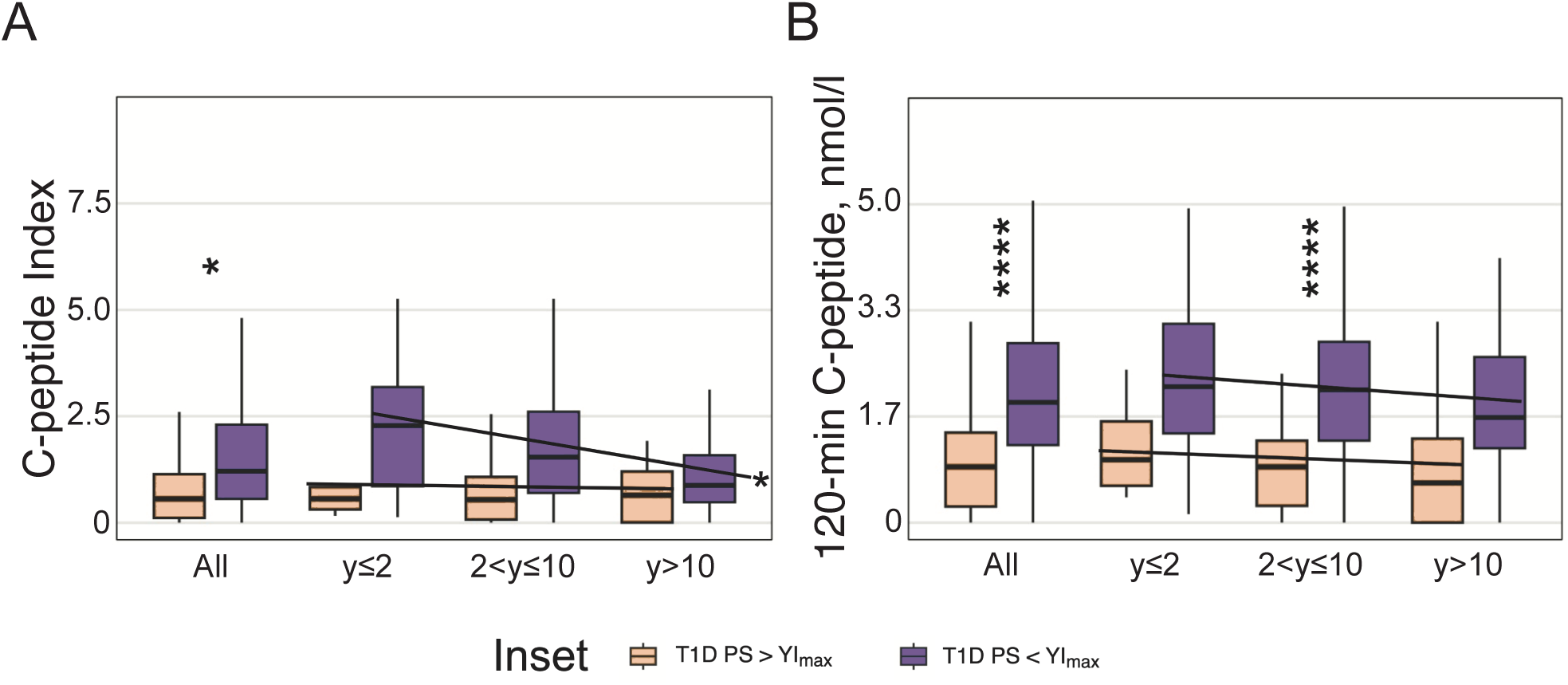
Relationship between T1D PS and C-peptide measures changes with diabetes duration. Boxplots of C-peptide measures obtained during OGTT, stratified by T1D PS categories, and plotted first for the entire RADIANT cohort (“All”) and then subset by duration of diabetes (y in years). Pale orange boxes show individuals with T1D PS>YI_max_ (12.88), while purple boxes show individuals with T1D PS<YI_max_. Boxplots show the median with interquartile range (IQR), with whiskers extending to 1.5x IQR. Mean C-peptide measures for T1D PS>YI_max_ versus T1D PS<YI_max_ were compared using linear regression models adjusting for standard covariates and meta-analyzed across ancestries. Trend lines reflect change in C-peptide measures over diabetes duration categories for each T1D PS category. Significance is labeled to the right side of the corresponding trend line with an asterisk. Significance surpassing Bonferroni correction is labeled with asterisks; *p*<0.05, *; *p*<0.0001, ****. **A**, C-peptide index. For T1D PS<YI_max_, p-trend slope *p*=0.0065*; T1D PS>YI_max_, p-trend slope does not reach significance. **B**, 120-min C-peptide (nmol/l). Neither p-trend slope reaches significance.

### Individuals with high T1D PS have 13-fold higher odds of a “type 1 diabetes-<u>like</u>” phenotype

Finally, we assessed whether the high-T1D PS group could identify individuals with a “type 1 diabetes-<u>like</u>” phenotype. Among 57 individuals with a high T1D PS and complete clinical data, 18 individuals (31.6%) met criteria for “type 1 diabetes-<u>like</u>” phenotype (Supplemental Table 2) compared to 12 of 244 other individuals (4.9%). In our adjusted, ancestry meta-analyzed models, those in the high T1D PS group had 13.2-fold higher odds of a “type 1 diabetes-<u>like</u>” phenotype compared to all other participants (Supplemental Fig. 3B). If a higher T1D PS threshold was used to define high T1D PS, we observed an even greater proportion of individuals with a “type 1 diabetes-<u>like</u>” phenotype, e.g. 50% at a score threshold of 15.19 (Supplemental Table 3).

Interestingly, among 40 individuals with high T1D PS and complete data who did not meet all criteria for a “type 1 diabetes-<u>like</u>” phenotype, 52.5% (21/40) required basal and bolus insulin or had low C-peptide levels, suggesting that T1D PS could be informative in these cases as well. Among those with high T1D PS, a “type 1 diabetes-<u>like</u>” phenotype, and complete data, 47% (8/17) had a history of DKA, and 50% (7/14) had a family history of autoimmunity.

Among 12 individuals who did not have high T1D PS but displayed a “type 1 diabetes-<u>like</u>” phenotype, 6/12 had a T1D PS above 11.5, which was recently proposed as an alternative threshold with improved sensitivity to detect type 1 diabetes [28]. This group was also enriched with individuals of non-European ancestry (5/12). However, these individuals of non-European ancestry with a “type 1 diabetes-<u>like</u>” phenotype still had a T1D PS below 11.5.

## Discussion

Using the largest and most comprehensively phenotyped cohort of individuals with IAb-unsolved diabetes, we demonstrate that diabetes polygenic scores aid in characterizing atypical diabetes and provide prognostic information where current clinical tools are insufficient. Despite attempting to exclude classic type 1 diabetes and type 2 diabetes, individuals in RADIANT had elevated T1D PS and T2D PS compared to individuals without diabetes in an ancestry-matched population. Stratifying individuals by T1D PS enabled identification of a high T1D PS group with OGTT measures indicative of reduced β-cell function, who were six times more likely to be on intensive insulin therapy and 13-times more likely to have a “type 1 diabetes-like” phenotype. A high T1D PS was thus strongly predictive of those who required intensive insulin therapy and had evidence on OGTT of reduced β-cell function. These findings demonstrate the potential clinical utility of the T1D PS in this population of IAb-atypical diabetes.

Specifically, since the T1D PS strongly predicted phenotypic features like insulin deficiency and need for both basal and bolus insulin, the T1D PS may be useful in individuals with IAb-atypical diabetes for flagging which patients should be monitored for insulin deficiency. While a high T1D PS may not equate to a diagnosis of type 1 diabetes, it could provide clinically-actionable information for clinicians. Failure to start insulin might lead to DKA and other complications [29]. In our study, some individuals with high T1D PS were not started on basal insulin after diagnosis and developed DKA, including one who received an SGLT-2 inhibitor. In retrospect, knowledge of the elevated T1D PS may have encouraged earlier consideration of insulin initiation, preventing DKA.

This study extends previous work undertaken in more typical, well-defined forms of diabetes. In youth-onset diabetes, T1D PS identified individuals most likely to progress to insulin deficiency in the IAb-subset of the SEARCH study [30]. In adults with IAb+ type 2 diabetes, those with a higher T1D PS progressed to insulin deficiency more rapidly [31]. However, the T1D PS did not predict an insulin requirement in IAb-individuals. Unlike these other scenarios, our work is the first to be dedicated entirely to unsolved, real-world, atypical diabetes cases, where additional reliable tools could provide benefit.

How best to utilize these PS remains an open question. In our study, combining T1D PS and T2D PS did not substantially change clinical predictions, compared to just using T1D PS alone. This finding is similar to previous studies in type 1 diabetes [6] and latent autoimmune diabetes in adults [32], but it contrasts with another approach that used both T1D PS and T2D PS together to predict phenotypes [33]. The T1D PS was more strongly associated with outcomes than the T2D PS, which is consistent with their reported discriminatory capacities for distinguishing respective cases from controls (AUC∼0.9 for T1D vs AUC∼0.7 for T2D) [5, 12]. Nevertheless, T2D PS was associated with several glycemic and clinical phenotypes, and its predictive ability may continue to increase with forthcoming larger and more diverse T2D GWAS [34].

T1D PS and T2D PS are not known to be correlated, but we assessed their correlation in our dataset given that the T2D PS results mirrored the T1D PS results, but in the opposite direction. We identified a non-significant negative correlation between T1D PS and T2D PS in RADIANT (Spearman ρ=-0.12, *p*=0.06 after meta-analysis), which differed from the weakly positive, very significant association seen among 292,400 controls in the UKBB of all ancestries (ρ=0.048, *p*=5.2×10^-148^). The differing T1D and T2D relationship in RADIANT could be influenced by collider bias, an artifactual association caused by conditioning on a shared outcome. Because T1D PS and T2D PS both increase diabetes risk, selecting for individuals with atypical diabetes in RADIANT could have induced the negative correlation between the two scores. Indeed, when we analyzed all individuals in the UKBB with any form of diabetes and selected for BMI 18.5–27 kg/m^2^, we observed a negative correlation between T1D PS and T2D PS (ρ=-0.09, *p*=7.2×10^-10^), which is likely consistent with collider bias.

Our data support the potential for T1D PS to improve the clinical care of atypical IAb-cases of diabetes. However, just like other pieces of clinical data to define diabetes type, T1D PS are not 100% sensitive or specific. Caution must be used in interpreting PS, especially between different ancestries [28]. Within RADIANT, the high T1D PS group had fewer individuals with African ancestry and none with East Asian ancestry. While the paucity of non-European participants with high T1D PS may reflect the underlying prevalence of type 1 diabetes or “type 1 diabetes-<u>like</u>” features in these populations, it is also known that this specific T1D PS (T1D GRS2) is significantly lower in individuals of African ancestry [28]. We attempted to alleviate this issue by controlling for genetic ancestry; however, the optimal T1D PS threshold may be different across populations. Alternatively, a T1D PS developed in individuals of African ancestry could be applied [35]. However, ancestry-specific scores are not available in all non-European populations. Recently developed scores have similar distributions across ancestries and may mitigate this challenge [36, 37]. If implemented correctly, PS could reduce disparities in diabetes diagnosis by ancestry [38].

While we explored the implementation of T1D PS and T2D PS in this study, there are now partitioned type 2 diabetes PS (pPS) [39], which assess for genetic variants in specific pathways thought to be driving disease phenotype [40]. Additional studies will be needed to understand the utility of pPS in those with atypical diabetes.

Strengths of this current study include the largest comprehensively phenotyped cohort for individuals judged to have “atypical” diabetes by a panel of experts. Therefore, this represents the best cohort to test implementation of PS in real-world “atypical” diabetes. There are a few limitations of this study. For genetics, the study size was small, and the study lacks longitudinal data. Some participants only had diabetes for a few months and others for over 20 years; it is possible that insulin requirements and islet autoantibody status may change over time, and some individuals with high T1D PS may progress to insulin deficiency with further follow-up. Additionally, while rapid-acting insulin was not administered during OGTT, other medications, including GLP-1 receptor agonists, could have affected measures of insulin secretion and other phenotypic variables.

In conclusion, we demonstrate that PS reveal genetic heterogeneity in the composition of individuals with atypical diabetes. Even though individuals were IAb-, we nevertheless identified a subset of individuals with high T1D PS who were enriched for treatment with insulin and reduced β-cell function. Given these findings, we postulate that PS could have clinical utility. New studies are needed to prepare these PS for clinical implementation.

## Data Availability

RADIANT has adopted policies and procedures in support of its commitment to sharing data with the scientific community while also protecting the privacy of participants.
RADIANT has established procedures to submit data to the NIDDK Central Repository in segments that reflect study progress. Access and availability of data submitted to the NIDDK Central Repository is determined by the NIDDK.
RADIANT has also established a policy for the release of RADIANT data not in the NIDDK Central Repository. Please refer to the RADIANT Policy on the Release of RADIANT Data and the Data and Materials Distribution Agreement (DMDA) for more information. Requests for access to data not in the NIDDK Central Repository may be submitted from the community of investigators currently working on RADIANT or from others who would like to collaborate with RADIANT in accordance with this policy and DMDA. Collaborators (i.e. researchers who are not RADIANT Study Investigators) who wish to access RADIANT study data need to team up with a RADIANT Study Investigator willing to sponsor their research as described in the DMDA.
To request access to RADIANT study data, please email the RADIANT DCC Team at

## Data Availability

The datasets generated during and/or analyzed during this study are not currently publicly available, but are available from the RADIANT study upon request, with more information provided on the RADIANT study website: https://www.atypicaldiabetesnetwork.org/researchers.

## Funding and Assistance

The RADIANT Study is funded by U54 DK118638 and U54 DK118612 from the National Institute of Diabetes and Digestive and Kidney Diseases (NIDDK). This publication was also made possible by K12DK133995 (David Maahs, Linda DiMeglio, Multi-Center Program Directors) from NIDDK Physician-Scientist Career Development Award (RJK); Pediatric Endocrine Society Fima Lifshitz Junior Faculty Clinical Research Grant (RJK); NIH NIDDK K23DK140643 (AJD); American Diabetes Association 11-23-PDF-35 (AH-C); NHLBI K24 HL157960 (JCF); American Diabetes Association 11-22-ICTSPM-16, NHGRI U01HG011723, NIDDK R01DK137993, R01DK140545, and U01 DK140757, a Medical University of Bialystok (MUB) grant from the Ministry of Science and Higher Education (Poland), and Novo Nordisk NNF21SA0072102 (JMM); Doris Duke Clinical Scientist Development Award 2022063 (MSU). The content is solely the responsibility of the authors and does not necessarily represent the official views of the National Institutes of Health.

## Authors’ Relationships and Activities

R.J.K. has consulted for mannkind, unrelated to this work. M.S.U. has provided consulting services for Novo Nordisk.

## Prior Presentation

Parts of this study were presented in abstract form at the American Society of Human Genetics Annual Meeting, October 14-18, 2025, Boston, Massachusetts and as a presentation at the 2026 American Diabetes Association Scientific Session, June 5-8, 2026, New Orleans, Louisiana. Parts of this study will also be presented at the Breakthrough T1D Clinical & Research Congress, October 9-11, 2026, Philadelphia, Pennsylvania.

## Figure Legends

**Supplemental Figure 1. Distribution of ADHD and Height PS among UK Biobank (UKBB) and RADIANT participants of European Ancestry. <u>A & B</u>** Violin plots of ADHD PS (**A**) or Height PS (**B**) distributions for UKBB and RADIANT participants of European ancestry. Significant differences in means were determined by Welch’s t-test; n.s., *p*>0.05. **C**, European participants’ ADHD PS sorted by rank. Black circles, RADIANT participants; blue circles, UKBB participants. **D**, European participants’ Height PS sorted by rank. Black circles, RADIANT participants; Blue circles, UKBB healthy control participants.

**Supplemental Figure 2. Interaction between T1D PS and T2D PS with C-peptide levels and clinical history. A and B,** Association between C-peptide measurements obtained during oral glucose tolerance test (OGTT) fit to linear models (left) or clinical history fit to logistic models (right), either including both T1D PS and T2D PS as covariates in the same model, or modeling T1D PS and T2D PS independently. Whether T2D PS and T1D PS are modeled together or independently, all models control for age, sex, BMI, diabetes duration, and 10 PCs of genetic ancestry.

Regression models were performed separately within each ancestry and then meta-analyzed. SD, standard deviation; OR, odds ratio. Significance surpassing Bonferroni correction is labeled with asterisks; *p*<0.01, **; *p*<0.001, ***; *p*<0.0001, ****. **A**, T2D PS. Blue triangles, T2D PS modeled without T1D PS as a covariate; pale orange circles, T2D PS modeled with T1D PS as a covariate. **B**, T1D PS. Purple triangles, T1D PS modeled without T2D PS as a covariate; red circles, T1D PS modeled with T2D PS as a covariate.

**Supplemental Figure 3. Association of C-peptide traits, diabetes prescription status, and participant history with T1D PS as a binary variable.** Participants were stratified into two categories depending on whether their T1D PS was greater than or less than 12.88 (the point with maximum Youden’s Index [YI_max_]). **A**, Association between C-peptide measurements obtained during oral glucose tolerance test (OGTT) and T1D PS. **B**, Association between participant history and T1D PS. Significance testing was performed with linear regression models (A) or logistic regression models (B) controlling for Age, Sex, BMI, duration, and 10 PCs of genetic ancestry. Regression models were performed separately within each ancestry and then meta-analyzed. *p*<0.05 from logistic and linear regression models are labeled. SD, standard deviation; OR, odds ratio. Significance surpassing Bonferroni correction is labeled with asterisks; *p*<0.01, **; *p*<0.001, ***; *p*<0.0001, ****.

**Supplemental Figure 4. T1D PS correlates with C-peptide levels.** Boxplots of C-peptide measurements obtained during OGTT, stratified by T1D PS, and plotted for the entire RADIANT cohort (“All”), or subset by duration of diabetes (subset y in years). Pale orange boxes show individuals with T1D PS > YI_max_ (12.88), while purple boxes show individuals with T1D PS < YI_max_. Boxplots show the median with interquartile range (IQR), with whiskers extending to 1.5x IQR. C-peptide levels for T1D PS > YI_max_ vs. T1D PS < YI_max_ were compared using linear regression models using the standard covariates within each ancestry and meta-analyzed. *p<*0.05 are labeled vertically.

Significance surpassing Bonferroni correction is labeled with asterisks; *p*<0.0001, ****. No p-trend slopes were significant after Bonferroni correction. **A**, Fasting C-peptide (nmol/l). **B**, 90-min C-peptide (nmol/l).

