## Supplemental Table 1 for "Diabetes Polygenic Scores Predict Glycemic Indices and Insulin Use in Individuals with Atypical Diabetes"

|  | T1D PS > YI_max_,  N = 59 | T1D PS < YI_max_,  N = 250 |  |  |
| --- | --- | --- | --- | --- |
| Clinical Measures | Median (1Q, 3Q) | Median (1Q, 3Q) | OR | *P* |
| Age at Stage 3 visit (years) | 38 (29, 48) | 49 (38, 61) |  | 0.98 |
| Age at Diabetes Diagnosis (years) | 26 (17, 35.5) | 33 (24, 45) |  | 0.52 |
| Diabetes Duration (years) | 7 (3,15) | 11 (4, 21) |  | 0.58 |
| BMI (kg/m^2^) | 22.8 (21.4, 25.0) | 24.1 (21.9, 28.3) |  | 0.92 |
| HbA1c (mmol/mol) HbA1c (%) | 46 (41, 54) 6.35 (5.93, 7.07) | 49 (42, 60) 6.6 (6.0, 7.6) |  | 0.48 |
| Sex | N (%) | N (%) |  | *P* |
| Female | 27 (45.8%) | 140 (56.0%) |  | 1.0 |
| Male | 32 (54.2%) | 110 (44.0%) |  |  |
| Inferred Genetic Ancestry^‡^ | N (%) | N (%) |  |  |
| Admixed American | 5 (8.5%) | 23 (9.2%) |  | 0.11 |
| African | 1 (1.7%) | 33 (13.2%) |  | 0.027 |
| East Asian | 0 (0%) | 17 (6.8%) |  | NA |
| European | 50 (84.7%) | 157 (62.8%) |  | 2.3 x 10^-4^* |
| South Asian | 3 (5.1%) | 20 (8.0%) |  | 0.098 |
| Diabetes (DM) Medication Status | N (%) | N (%) |  |  |
| Any DM Medication | 57 (99.6%) | 239 (95.6%) | 1.4 | 0.79 |
| Insulin Prescription | 45 (76.3%) | 136 (54.4%) | 4.9 | 0.0011* |
| Basal Insulin Only | 4 (6.8%) | 22 (8.8%) | 0.82 | 0.81 |
| Basal + Bolus Insulin | 38 (64.4%) | 100 (40.0%) | 6.3 | 4.2 x 10^-5^* |
| Non-insulin DM Med | 30 (50.8%) | 184 (73.6%) | 0.38 | 0.024 |

**Supplemental Table 1. Clinical measures of RADIANT participants stratified by T1D PS as a binary variable.** Clinical measures of IAb- individuals with atypical diabetes in the current study. A subset of participants was classified as having high T1D PS, defined as greater than 12.88 (the point with maximal Youden’s Index [YI_max_]). Clinical measures and prescription status were determined during Stage 3 site visit. Continuous measures are presented as median values (1^st^ quartile, 3^rd^ quartile in parentheses). Significance (*P*) and odds ratios (OR) were determined by linear or logistic regression models using the following covariates: Sex, BMI, Age at Stage 3 visit, Duration (Age at Stage 3 visit - Age of Diabetes Diagnosis), and 10 PCs of genetic ancestry. If the outcome measure was one of the listed covariates, or was used to calculate a covariate, then that measure was not included as a covariate in the regression model. Regression models were performed separately in each ancestry and then meta-analyzed, unless subset by ancestry. *Statistically significant after Bonferroni correction. ^‡^Genetic ancestry was inferred by principal component analysis.
