## Supplemental Table 2 for "Diabetes Polygenic Scores Predict Glycemic Indices and Insulin Use in Individuals with Atypical Diabetes"

| ID | Sex | Age Group at Diagnosis (years) | Duration (years) | DKA^†^ | BMI (kg/m2) | Highest HbA1c (%) | Autoimmune Family Hx^†^ | T1D PS | Population  Percentile | T1D  Percentile | C-peptide Index |
| --- | --- | --- | --- | --- | --- | --- | --- | --- | --- | --- | --- |
| 1 | M | 36-40 | 5 | N | 21.7 | 12 | Y | 16.6523 | 0.9983 | 0.8996 | 0.03 |
| 2 | F | 11-15 | 49 | Y | 19.4 | 9 | Y | 16.6023 | 0.9981 | 0.8917 | 0 |
| 3 | M | 0-5 | 38 | N | 25.1 | NA | Y | 16.4237 | 0.9974 | 0.8622 | 0 |
| 4 | F | 6-10 | 10 | N | 26.2 | 11 | Y | 15.311 | 0.9855 | 0.6319 | 0.03 |
| 5 | M | 31-35 | 9 | Y | 31.7 | 13 | N | 15.2288 | 0.984 | 0.6122 | 0.19 |
| 6 | M | 21-25 | 7 | N | 20.8 | 8.7 | NA | 14.8923 | 0.9765 | 0.5099 | 0.21 |
| 7 | F | 11-15 | 4 | Y | 20.5 | 14 | Y | 14.5062 | 0.9643 | 0.4311 | 0 |
| 8 | M | 21-25 | 15 | NA | 24.3 | 12.3 | Y | 14.4623 | 0.9627 | 0.4272 | 0.02 |
| 9 | M | 16-20 | 5 | N | 25 | NA | N | 14.3837 | 0.9596 | 0.4075 | 0.06 |
| 10 | M | 26-30 | 12 | N | 24.3 | 13.9 | N | 14.3748 | 0.9592 | 0.4075 | 0 |
| 11 | M | 51-55 | 48 | Y | 23.4 | 13.3 | N | 14.1668 | 0.95 | 0.3799 | 0.11 |
| 12 | F | 11-15 | 31 | Y | 20.5 | 13.1 | Y | 13.9588 | 0.9392 | 0.3189 | 0 |
| 13 | F | 31-35 | 9 | Y | 21 | 12 | NA | 13.6348 | 0.9189 | 0.2598 | 0.47 |
| 14 | F | 51-55 | 2 | N | 19.3 | 14 | NA | 13.6288 | 0.9184 | 0.2579 | 0.17 |
| 15 | F | 26-30 | 3 | Y | 22.3 | 12 | N | 13.2453 | 0.8883 | 0.1929 | 0 |
| 16 | M | 26-30 | 12 | N | 30 | 11.9 | N | 13.1648 | 0.8811 | 0.1772 | 0.12 |
| 17 | M | 26-30 | 5 | Y | 22.3 | NA | NA | 13.0212 | 0.8673 | 0.1516 | 0 |
| 18 | F | 16-20 | 25 | N | 23 | 7.5 | N | 12.9437 | 0.8596 | 0.1457 | 0 |

**Supplemental Table 2. Islet autoantibody-negative individuals from RADIANT with high T1D PS and “Type 1 Diabetes-Like” phenotype.** Clinical characteristics of individuals with T1D PS >12.88, islet autoantibody-negative, maturity-onset diabetes of the young (MODY) genetic testing negative, receiving either basal & bolus insulin or using pump, fasting C-peptide <0.20 nmol/l, and oral glucose tolerance test 120-min C-peptide <0.60 nmol/l. Age of diabetes diagnosis is presented in age group bucket in years. ^†^ Presence or absence of diabetic ketoacidosis (DKA) and autoimmune family history (Hx) are listed as Yes (Y), No (No), or Not Available (NA).
