## Supplemental Table 3 for "Diabetes Polygenic Scores Predict Glycemic Indices and Insulin Use in Individuals with Atypical Diabetes"

| T1D PS Threshold | Percentile Among  All Individuals in the UKBB | Percentile Among Individuals with Type 1 Diabetes  in the UKBB | Percentage of Individuals with “Type 1 Diabetes-Like” Phenotype Above Threshold in RADIANT (%)^†^ |
| --- | --- | --- | --- |
| 11.01 | 60.0 | 3.3 | 19.5 |
| 12.88 | 85.3 | 13.8 | 31.6 |
| 15.19 | 98.3 | 60.0 | 50.0 |

**Supplemental Table 3. Percentage of individuals with a “type 1 diabetes-like” phenotype above various thresholds of T1D PS.** A “type 1 diabetes-like” phenotype was assessed in 309 pancreatic autoantibody-negative RADIANT participants of all ancestries. Each row displays the percentage of individuals with a T1D PS above the threshold who had a “type 1 diabetes-like” phenotype. Total number of “type 1 diabetes-like” cases per ancestry: African, 2; Admixed American, 2; Asian, 1; European, 25. ^†^Percent calculated from raw T1D PS without covariates.
