## Supplementary figures and images for "Diabetes Polygenic Scores Predict Glycemic Indices and Insulin Use in Individuals with Atypical Diabetes"

### Supplemental Fig. 1

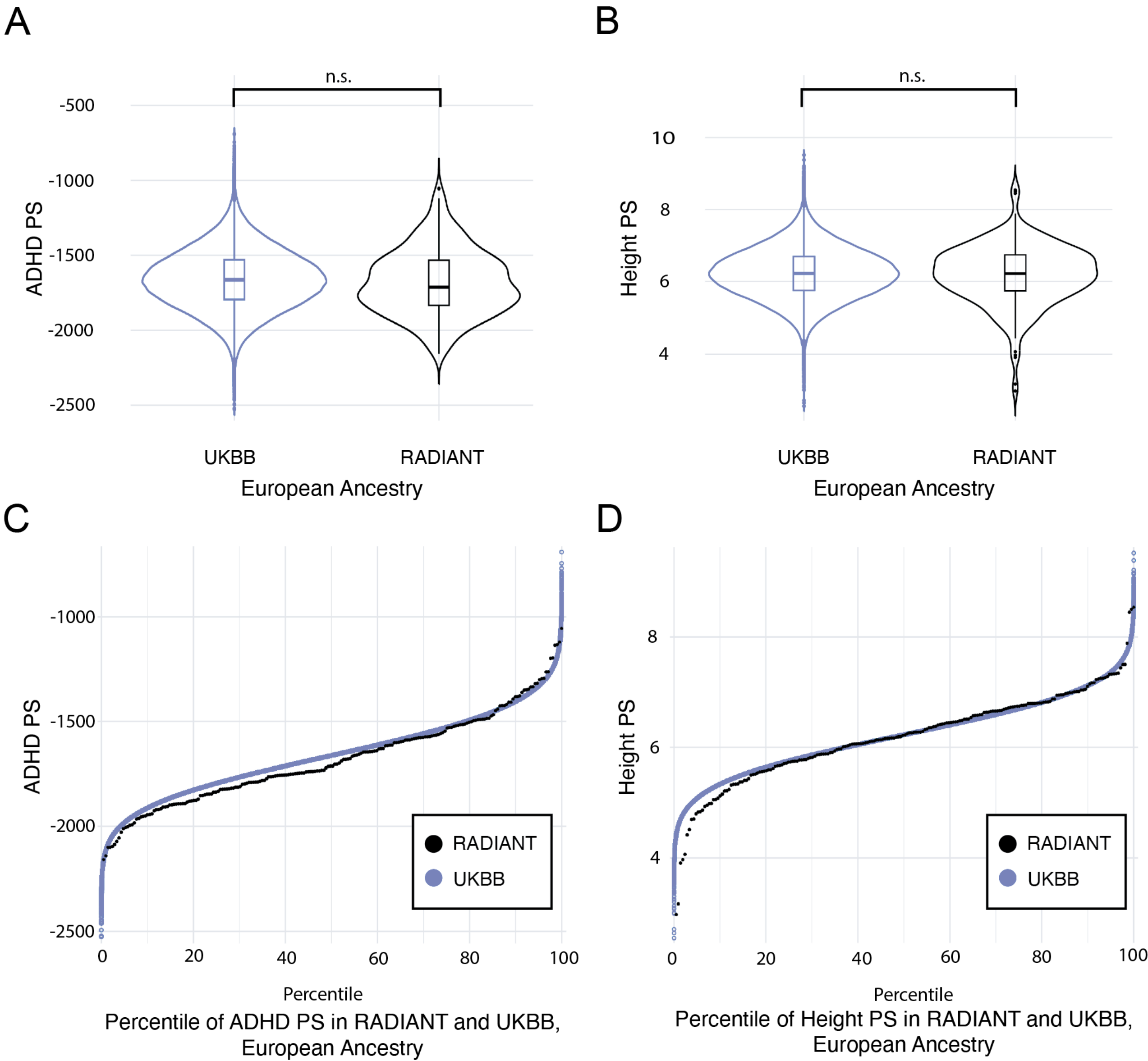

### Supplemental Fig. 2

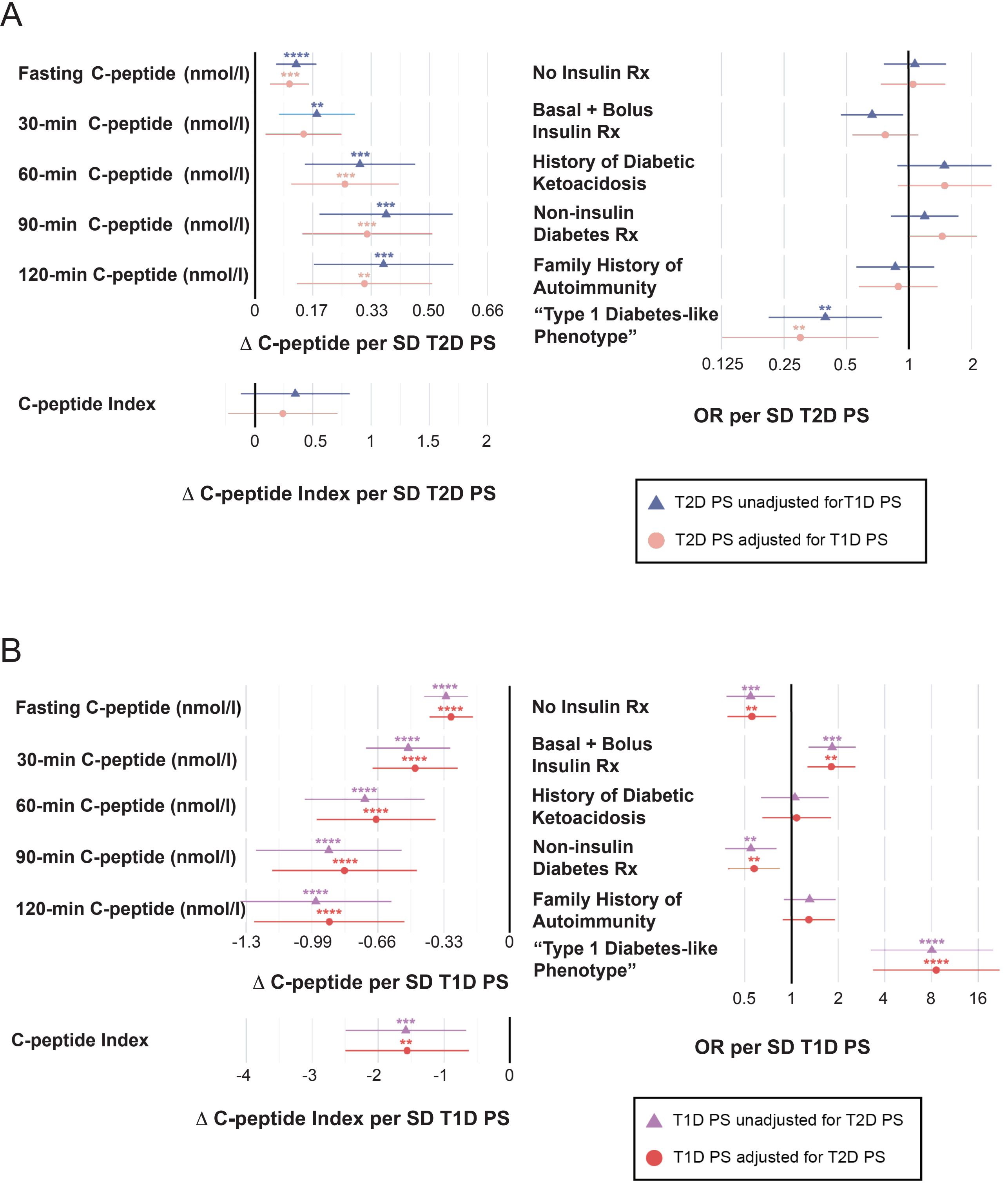

### Supplemental Fig. 3

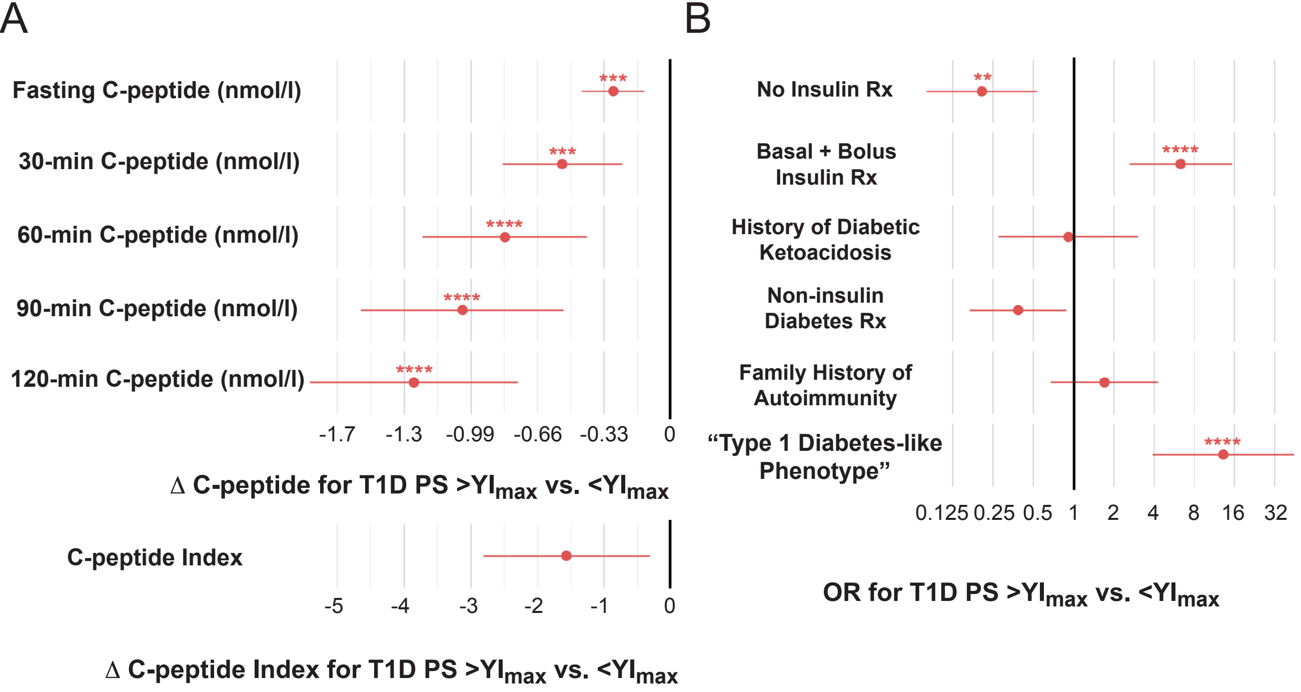

### Supplemental Fig. 4

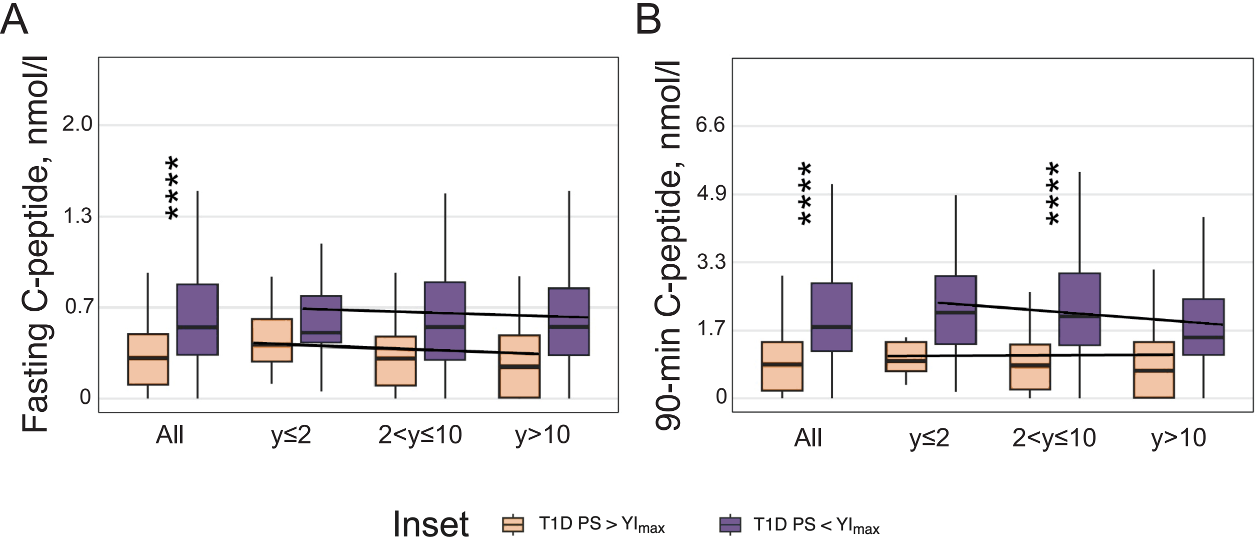
